# Risk of sustained SARS-CoV-2 transmission in Queensland, Australia

**DOI:** 10.1101/2021.06.08.21258599

**Authors:** Paula Sanz-Leon, Nathan J. Stevenson, Robyn M. Stuart, Romesh G. Abeysuriya, James C. Pang, Stephen B. Lambert, Cliff C. Kerr, James A. Roberts

## Abstract

We used an agent-based model *Covasim* to assess the risk of sustained community transmission of SARS-CoV-2/COVID-19 in Queensland (Australia) in the presence of high-transmission variants of the virus. The model was calibrated using the demographics, policies, and interventions implemented in the state. Then, using the calibrated model, we simulated possible epidemic trajectories that could eventuate due to leakage of infected cases with high-transmission variants, during a period without recorded cases of locally acquired infections, known in Australian settings as “zero community transmission”. We also examined how the threat of new variants reduces given a range of vaccination levels. Specifically, the model calibration covered the first-wave period from early March 2020 to May 2020. Predicted epidemic trajectories were simulated from early February 2021 to late March 2021. Our simulations showed that one infected agent with the ancestral (A.2.2) variant has a 14% chance of crossing a threshold of sustained community transmission (SCT) (i.e., *>* 5 infections per day, more than 3 days in a row), assuming no change in the prevailing preventative and counteracting policies. However, one agent carrying the alpha (B.1.1.7) variant has a 43% chance of crossing the same threshold; a threefold increase with respect to the ancestral strain; while, one agent carrying the delta (B.1.617.2) variant has a 60% chance of the same threshold, a fourfold increase with respect to the ancestral strain. The delta variant is 50% more likely to trigger SCT than the alpha variant. Doubling the average number of daily tests from ∼ 6,000 to 12,000 results in a decrease of this SCT probability from 43% to 33% for the alpha variant. However, if the delta variant is circulating we would need an average of 100,000 daily tests to achieve a similar decrease in SCT risk. Further, achieving a full-vaccination coverage of 70% of the adult population, with a vaccine with 70% effectiveness against infection, would decrease the probability of SCT from a single seed of alpha from 43% to 20%, on par with the ancestral strain in a naive population. In contrast, for the same vaccine coverage and same effectiveness, the probability of SCT from a single seed of delta would decrease from 62% to 48%, a risk slightly above the alpha variant in a naive population. Our results demonstrate that the introduction of even a small number of people infected with high-transmission variants dramatically increases the probability of sustained community transmission in Queensland. Until very high vaccine coverage is achieved, a swift implementation of policies and interventions, together with high quarantine adherence rates, will be required to minimise the probability of sustained community transmission.

## 1. Introduction

The novel coronavirus (COVID-19) pandemic has altered normal daily life for the majority of the world’s population, tallying over 332 million confirmed cases and over 5.5 million deaths globally (https://covid19.who.int/ – accessed January 20th 2022). A key area of COVID-19 research has been the development of models that capture the spread of the virus under a wide variety of conditions and incorporate demographics of specific geographic locations [1–8]. These models have been used to simulate interventions such as ‘stay at home’ orders, school closures, social distancing, border closures, and vaccination. Analyses of simulation outputs provide insights on the efficacy of interventions, permit stress testing of a range of interventions against increasing infection rates, and help estimate the use of resources required for desired health outcomes. This information can guide policy makers to find the delicate balance between minimising the spread of COVID-19, and increasing social and economic disruption.

In Australia, following the closure of international borders in March 2020, much of the response to the pandemic has been undertaken by state and territory governments. In the Australian context, local elimination of COVID-19 transmission has occurred through the implementation of location-specific interventions. To accurately simulate this scenario, state-specific apparent transmissibility (the probability that an infected person will transmit COVID-19 to another), the age distribution of the population, and differences in lockdown policies must be considered.

The assessment of existing policy effectiveness against the transmission of mutated strains of the virus is of particular relevance, with numerous variants emerging overseas [9]. One variant of concern is the B.1.1.7 (or alpha) lineage that emerged in the UK. It is estimated to be between ∼ 40%–90% more transmissible than previous COVID-19 strains, though there is no apparent change in disease severity (e.g., critical cases and deaths) [10]. Variants such as the B.1.617.1 (or kappa) and B.1.617.2 (or delta) lineages that emerged in India have been shown to be up to 2.5 times more transmissible than the ancestral strain [9, 11–13]. In countries with ongoing community transmission, more transmissible variants rapidly dominate. However, in Queensland, a setting with prolonged periods without recorded cases of locally acquired infections (i.e., zero community transmission), the effects of new variants with increased transmissibility on the spread of COVID-19 remain poorly understood [9], despite the fact that other Australian states experienced large outbreaks of the Delta variant.

In this paper, we calibrate an agent-based model of COVID-19 transmission (Covasim [14]) to epidemiological data in Queensland, Australia, taking into account the state’s mix of policies and the timing of their implementation. This model complements existing Covasim models for the spread of COVID-19 in other Australian states including New South Wales [15] and Victoria [16].

We use the calibrated model to estimate the probabilities of sustained community transmission regimes. We compare the spread of COVID-19 with (a) the ancestral strain (A.2.2 in the PANGO nomenclature [17]); (b) the alpha variant [17]; and, (c) the delta variant, in a Queensland setting. The role of testing rate as a mitigating intervention is also investigated.

## 2. Methods

The model used a population of 200,000 agents to predict the transmission of COVID-19 in Queensland, generated with the demographic data from the 2016 census by the Australian Bureau of Statistics [18]. Agents can interact deterministically (e.g., agents who meet regularly at places of worship) or randomly (e.g., entertainment venues, festivals, exhibitions). The probability that an agent is infected depends on interactions with their contact networks; here we modelled 14 different networks [14] (see Supplementary Fig. S1). Other variables were set according to current literature or modified for the Australian milieu [14, 16].

Start and end dates of policies and interventions were compiled from multiple online sources [19, 20] (Supplementary Fig. S2). The effect of policies on the spread of COVID-19 were modelled as a network-specific reduction in transmissibility. We used previous Australia-specific Covasim models [16] to estimate the effect of policy changes in each contact network (Supplementary Fig. S1 built from multiple sources [19–21]), as well as other parameters that were not inferred during the calibration process (Supplementary Table S1).

The model parameters inferred from Queensland-specific data were: (i) overall transmissibility; (ii) initial number of infections on 1 March 2020; and (iii) testing odds ratio (i.e., how much more symptomatic agents are tested compared to asymptomatic agents). The testing odds ratio is adjusted over three critical periods in 2020: 1 March to 29 March, 30 March to 16 April, and 17 April to 15 May (date after which locally transmitted cases reached zero). The data used to calibrate the model were acquired directly from the Epidemiology Team at Queensland Health and included the number of infected cases (total, acquired locally, and acquired overseas), and tests per day, with data spanning back to Queensland’s first confirmed case on 28 January 2020.

A two-stage process was undertaken to fit the model. Both steps minimised the absolute differences between the median model trajectory and the empirical trajectory of locally-acquired, cumulative diagnoses. The first step used a fixed-grid search to determine suitable ranges for the model parameters. The second step employed a hyperparameter optimisation framework [22] to adjust five parameters simultaneously (overall transmissibility, initial number of infections on 1 March 2020, and the testing odds ratio in the three periods mentioned above). The simulated period spanned 1 March to 15 May 2020. Samples of parameter combinations were drawn from a uniform distribution. For all simulations during the fitting process, we incorporated the empirical daily number of tests administered during that period and assumed a high level of compliance when agents are instructed to quarantine at their home, similar to other Australian settings with very few locally acquired cases that can be easily contained with such interventions (also known as low-community transmission settings) [15].

Using the calibrated model, we then explored the impact of “introduction scenarios” in which infectious agents were released into the community. In these scenarios we used policies and/or behavioral changes representative of the period from late January 2021 to late June 2021, collectively known as *the new normal*. These were social distancing (1.5 m), staying at home when sick, non-mandatory mask wearing (i.e., wearing mask when distancing was not possible), and increased hand hygiene [23]. We did not include the 3-day lockdown that took place in the middle of March 2021 because the purpose of these scenarios was to assess the susceptibility of the population in the context of the “new normal”. All scenarios included contact tracing. The average number of days and probabilities of being traced for each layer can be found in Supplementary Table S2.

The main output measure in these introduction scenarios was the probability of crossing a threshold that would indicate sustained community transmission (SCT) leading to a potential outbreak. SCT was defined as occurring when the number of *new infections* is greater than 5 per day for at least 3 consecutive days. We also measured the probability of detecting SCT (DSCT), using *new diagnoses*, which are the observable upon which policy changes would be enacted. Specifically, DSCT occurs when the 3-day average of *new diagnoses* is greater than 5 per day for at least 3 consecutive days. Both definitions are based on a modelling study of outbreak projections for Victoria [24]. Probabilities were quantified as the percentage of model simulations that crossed the corresponding threshold (SCT or DSCT), out of 1000 simulations.

All epidemic projections span the period from 1 February 2021 to 31 March 2021. We seeded a *cluster* of infected agents into the community by infecting a subset of agents in a single day. The agents within the cluster are assumed to be unlinked (i.e., they do not belong to the same household or workplace). This approach models scenarios such as an undetected leak from hotel quarantine, non-compliance of at-home quarantine instructions, or undetected clusters in the community. Since March 2020, in response to the global COVID-19 situation, people arriving in Australia from overseas or a national COVID hotspot are required to quarantine in government arranged accommodation (e.g., a hotel). This is known as hotel quarantine.

We simulated three main scenarios: (i) a cluster of seeded infections for the current calibrated Queensland transmissibility; (ii) a cluster of seeded infections with the transmissibility increased by 70% to simulate a alpha-like variant; and, (iii) cluster of seeded infections with the transmissibility increased by 150% to simulate a delta-like variant.

We repeated these analyses for a stream of incoming infections (termed *Poisson* seeding), by releasing a small number of infected agents randomly (in number and time) over the simulated period. This approach incorporates the random arrival time of infected agents and could model international travel bubble scenarios, or repeated failures of hotel quarantine with a reservoir of active cases (Supplementary Figs S3–S5). For each scenario, each model run uses a different realisation of the initial cluster (or stream) of infections.

We examined the mitigating effects on SCT of *testing rate*, and subsequent at-home quarantine/isolation (Q&I). Here, we define at-home quarantine as segregation of potentially infected agents (not to be confused with hotel quarantine) and (at-home) isolation as segregation of confirmed (via testing) infected agents. When infected agents are detected via testing, they are put in isolation (I). As a secondary effect the contacts of confirmed cases are traced and tested (with higher priority than the general population). The exposed contacts are then quarantined (Q) while they wait for their test result to come back. The effect of testing was simulated by varying the daily number of tests and Q&I leakage. We allocated tests randomly throughout the population. The minimum baseline number of tests was estimated from empirical data as the mean number of daily tests between 22 January and 22 February 2021 (6260 ± 2100 tests, mean ± SD), though in the Results section we present several results with ∼ 8,000 daily tests, a number that better reflects empirical data from May-June 2021 (https://covidlive.org/ – accessed September 29th 2021) . The effects of Q&I were simulated by varying “leakage” (non-adherence) from quarantine and isolation. The lower these values, the fewer infected agents (potential and confirmed) re-enter the community.

Finally, we also investigated the effects of combinations of *vaccination effectiveness* and *vaccination coverage*. In this work, vaccine effectiveness values correspond to the prospective level of protection against infection after completing the recommended vaccine treatment [25]. For instance, a vaccine effectiveness of 50% corresponds to halving the agent’s susceptibility to becoming infected. Vaccine coverage represents the percentage of people aged 18 and over, who have received the vaccine treatment.

## 3. Results

### Calibration Results

The calibrated model accurately captures the initial outbreak and the decline in cases following lockdown (Figure 1). The model also predicts that for every diagnosed case there were approximately four undetected infections in the community. The inferred model parameters suggest that Queensland was in a low community transmission setting in the period of March-May 2020.

**Figure 1.**
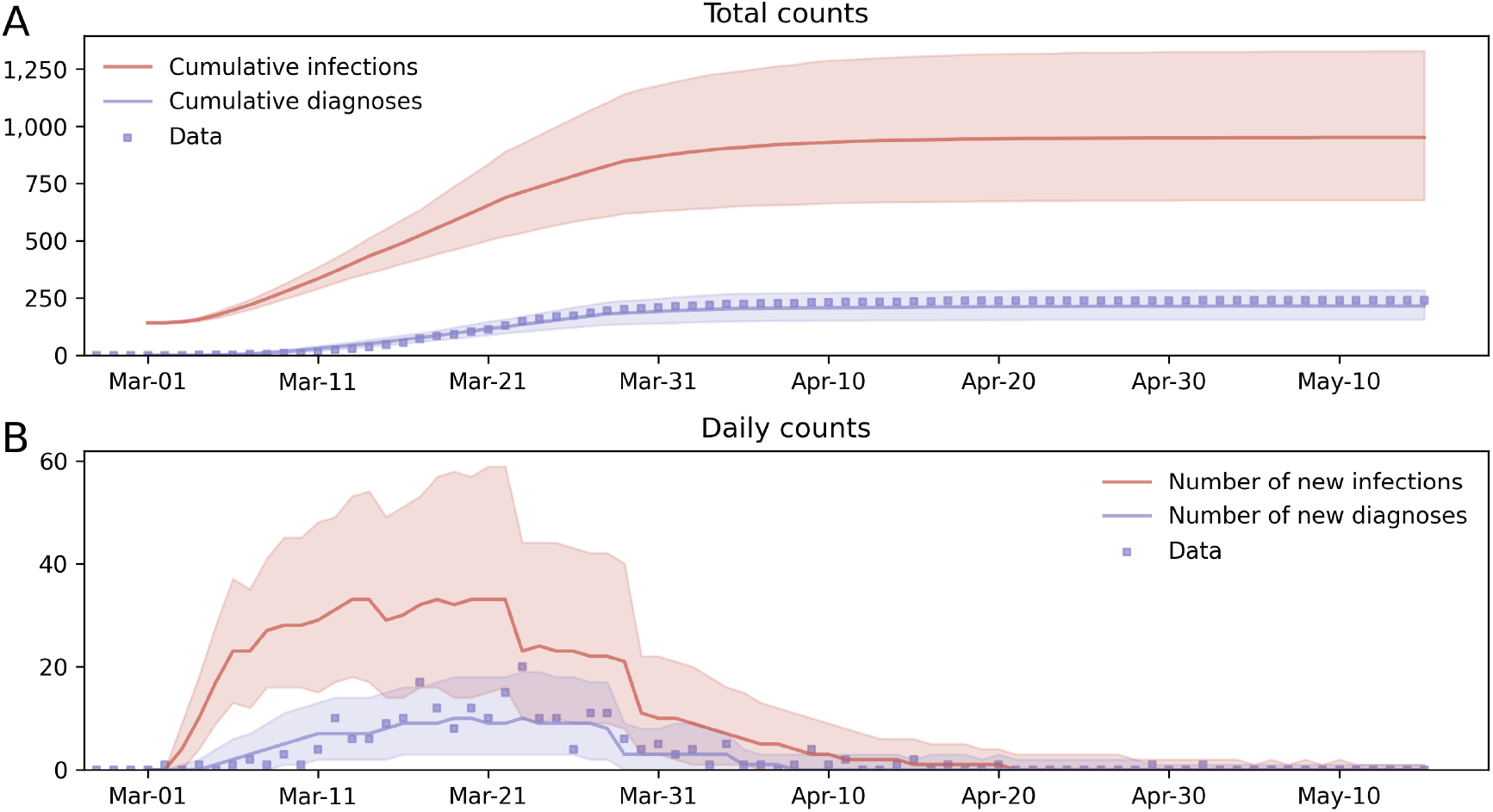
Fitting of the calibrated model to the main COVID-19 wave in Queensland between 1 March 2020 and 15 May 2020. **A**: Cumulative trajectories. **B**: Daily trajectories. Solid lines are the median model projections over 1000 model runs; shaded areas indicate 95% projected intervals over different initialisations of infected agents on 1 March 2020; blue dots represent empirical numbers of confirmed locally-acquired cases.

### Sustained Community Transmission Results

Using the calibrated model, we explored the susceptibility of the Queensland population to developing SCT given a cluster of seeded infections. Figures 2A-C illustrate the projected trajectories of new infections for a range of cluster sizes. The larger clusters generate higher daily infection rates more quickly, regardless of the variant. However, the ancestral variant exhibits markedly lower growth rates than the alpha and delta variants, with many instances of small initial clusters failing to cross the SCT threshold within the modelled 8-week period.

**Figure 2.**
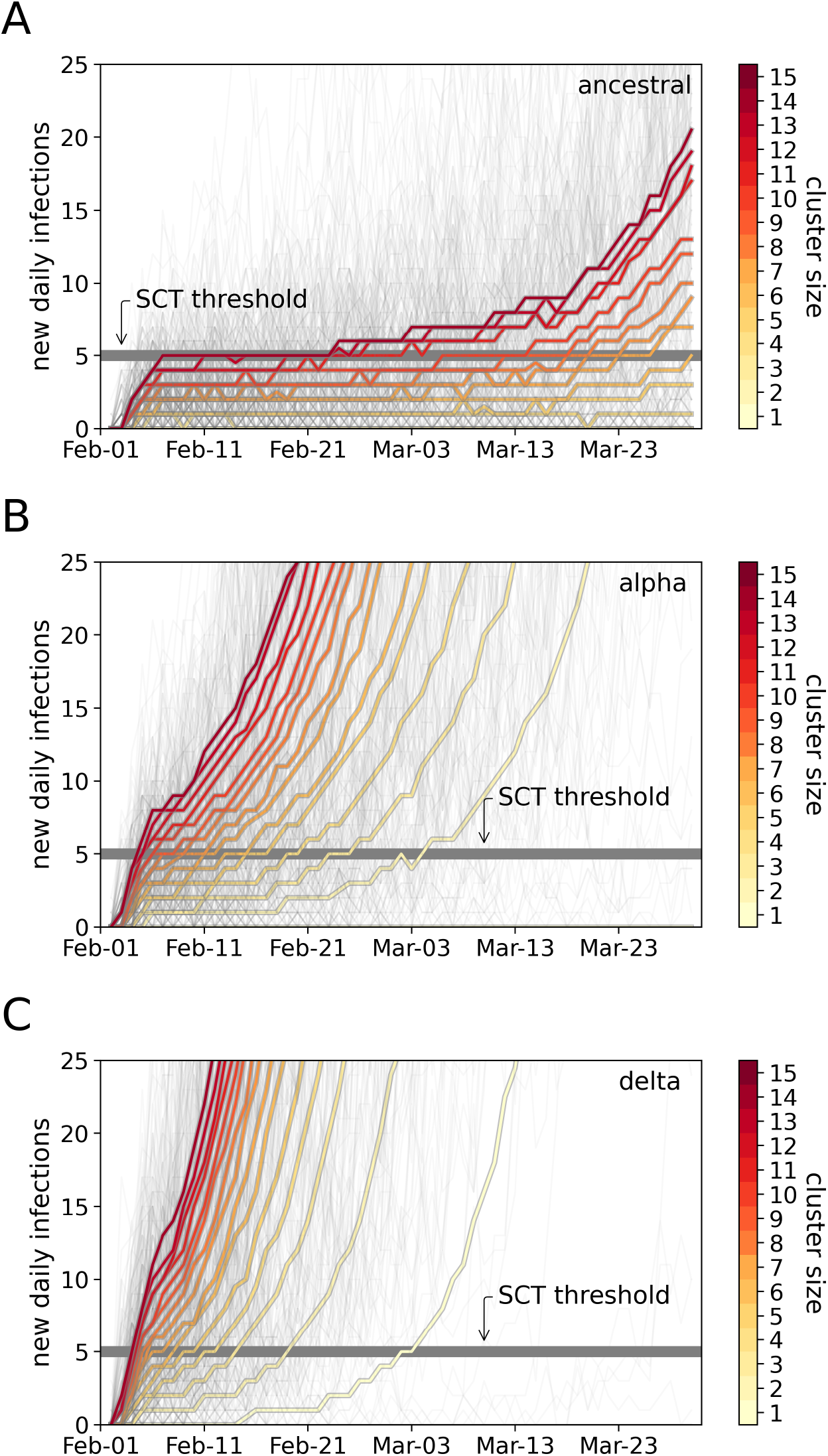
Predicted epidemic trajectories for new clusters of infections comparing the ancestral and alpha variants. **A:** Projected trajectories of new infections assuming Queensland-specific ancestral transmissibility and a range of seeded cluster sizes. **B:** Projected trajectories for the Queensland-specific alpha transmissibility. **C:** Projected trajectories for the Queensland-specific delta transmissibility. In A, B and C, thick coloured lines represent the median trajectory of new infections, estimated over 1000 runs. The colour of each line denotes the size of the initial cluster of infections. Light-grey thin lines are selected individual trajectories from scenarios with different cluster size.

We note that the almost linear increases in daily infections in the first 5 days of the median trajectories (coloured lines in Fig. 2) are a side effect of the seeding approach used in these scenarios. The number of daily infections immediately after inserting the cluster of infections on the first day of the simulations is proportional to the cluster size. The new infected agents consist of the immediate contacts of the infected agents in the cluster. After this short period the virus spreads throughout the whole community and the median trajectories exhibit the stereotypical exponential growth seen in large outbreaks.

Similar increasing growth rates for highly-transmissible variants are observed for streams of randomly-arriving (Poisson seeded) infections (Supplementary Fig. S3). We note that using this alternative seeding approach the almost linear increase in daily infections in the first 5 days is no longer present.

The probability of developing SCT is shown in Fig. 3A. For large clusters, all variants are likely to pass the threshold of SCT. However, for small clusters, the alpha and delta variants present a substantially higher risk of developing SCT. This result has only weak dependence on the rates of Q&I leakage (Supplementary Fig. S4). The Queensland-specific relative risk of SCT for the alpha and delta variants over the ancestral variant is shown in Fig. 3B. This ratio shows that for a single case, the probability of developing SCT in the presence of a variant that is 70% more transmissible than ancestral, is 300% higher than the probability of developing SCT in the presence of ancestral; and, the probability of developing SCT in the presence of delta is approximately 450% higher. The probabilities of DSCT are also strongly elevated (Supplementary Fig. S5).

**Figure 3.**
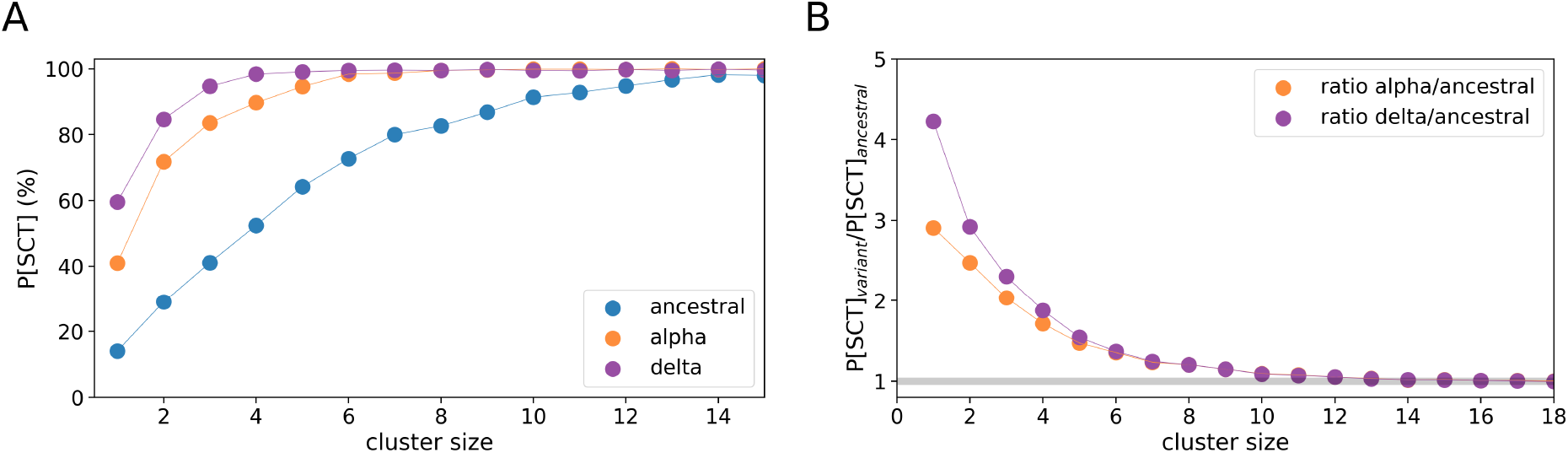
Probabilities of sustained community transmission (SCT) for different variants. **A:** Summary of probabilities of SCT for the ancestral variant (blue), alpha variant (orange) and delta variant (violet). **B:** Relative risk of SCT for alpha versus ancestral (orange), and for delta versus ancestral (violet) estimated as the SCT probability for a highly-transmissible variant divided by the SCT probability for ancestral. The grey line denotes when the SCT probability for both variants is the same. Simulations used a level of testing ∼8k daily tests and a moderate Q&I leakage (∼50%).

We also estimated how often it happens that a cluster dies out again after SCT was reached. For scenarios with ancestral strain clusters, this happens for approximately between 3–12% of the simulations that reached SCT. For scenarios with alpha and delta clusters, this happens for less than 1% of simulations that reached SCT. Further, in Poisson-seeded scenarios with the ancestral strain, less than 3% of simulations die off, while scenarios with the alpha and delta variants, less than 1% of simulations die off.

Next, we examined the effects of increased testing on the probabilities of developing SCT. Since increased testing leads to increased Q&I, it is plausible that this intervention could mitigate the increased transmissibility of new variants. Thus, we simulated a range of increased testing rates and calculated the ensuing probabilities of developing SCT (Fig. 4A). We found that doubling the level of testing, from ∼ 6k tests to ∼ 12k daily tests, results in a 23% decrease in the probability of developing SCT due to a single seeded infection of the alpha variant (i.e., a drop from approximately 43% to 33%). However, to achieve the same 23% decrease due to a single seeded infection of the delta variant, we would need to increase daily testing by an order of magnitude (10-fold). Even if we achieved this reduction of 23%, the risk of developing SCT due to a single infected agent (with either alpha or delta) leaking into the Queensland community would still remain approximately 2.5 times larger than the risk estimated for the ancestral variant. The 10-fold increase in testing to reduce the excess risk posed by high-transmission variants, while achievable is not sustainable in the long term.

**Figure 4.**
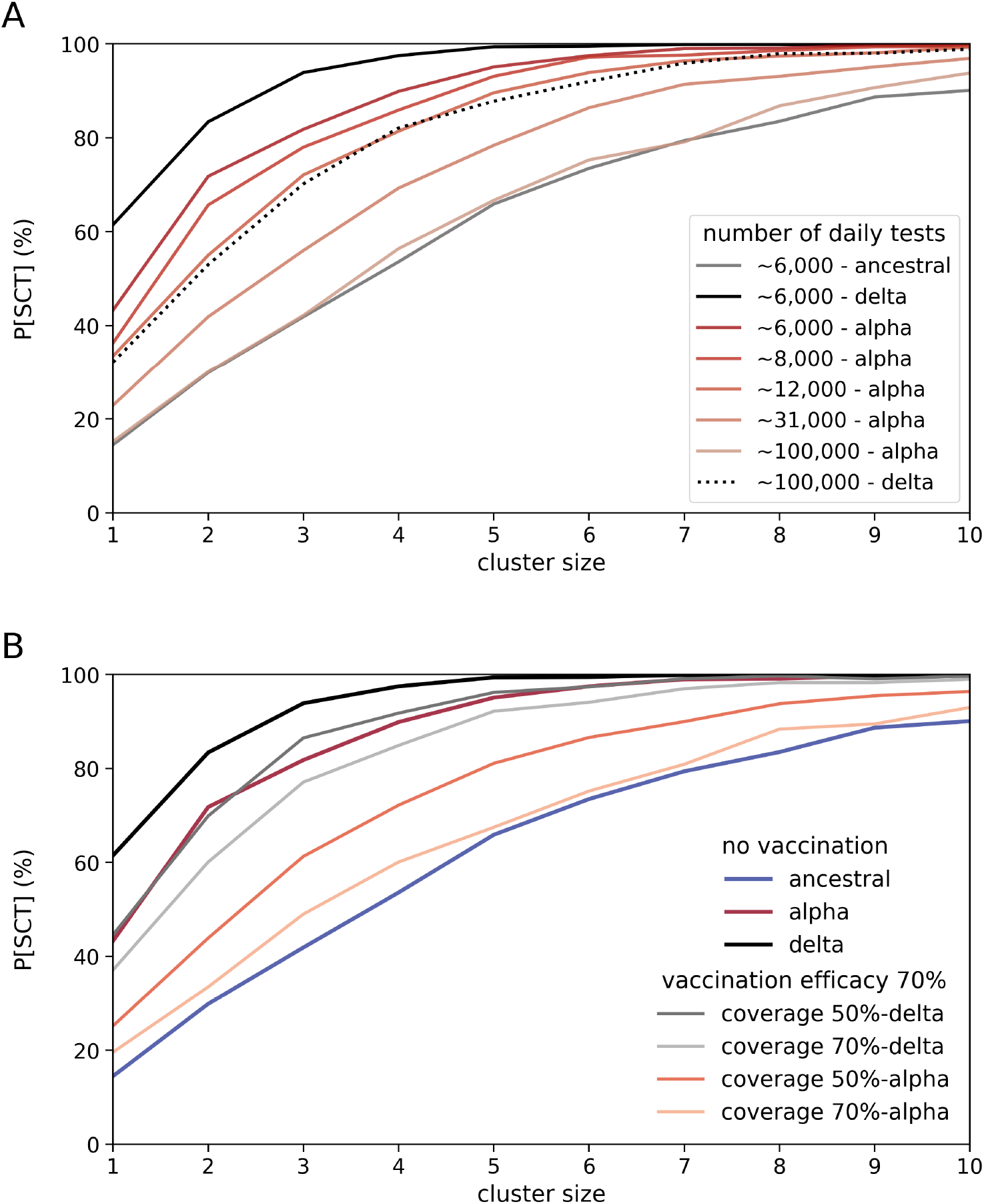
Effects of testing and vaccinations on the probabilities of sustained community transmission (SCT). **A:** Probabilities of SCT for multiple levels of testing. The dark grey line represents SCT probabilities for ancestral and ∼ 6000 daily tests. The black lines represents SCT probabilities for delta, for ∼ 6000 daily tests (solid line) and ∼ 100,000 daily tests (dotted line). Red-shaded lines are SCT probabilities for the alpha variant and multiple levels of testing. **B:** Probabilities of SCT for multiple combinations of variant and vaccine coverage, for a vaccine with 70% effectiveness against transmission. The black, dark red, and dark blue lines represent SCT probabilities for delta, alpha and ancestral, respectively, assuming ∼ 6000 daily tests, and no vaccination. The other solid lines are SCT probabilities as a function of vaccine coverage. Gray line 50% vaccinated and delta; light gray line 70% vaccinated and delta; red line 50% vaccinated and alpha; and, light red line 70% vaccinated and alpha. In these scenarios with vaccination, the cluster size represents the number of infections seeded on the first day of the simulated period. All cases have a low Q&I leakage (<10%).

We note that there is a two-sided effect in regards to increased testing and the behavior of the probability of DSCT. If we increase testing, more cases are found and DSCT may exhibit an increase. However, a sufficiently large increase in daily tests (e.g., ∼ 100,000) eventually leads to a decrease in the probability of DSCT (Supplementary Fig. S6) because of the active preventative role of testing via Q&I. When testing rate is sufficiently high, the likelihood of developing SCT decreases, to a point where SCT occurs so rarely that the likelihood of DSCT also decreases. This two-sided effect, and the difference between SCT and DSCT probabilities, stem from the finite number of tests carried out every day, and from the fact that our measurement instrument (i.e., testing) interferes with the phenomenon being measured (i.e., propagation of disease within the community) via secondary mechanisms (i.e., Q&I and tracing).

Finally, we examined the effects of increased vaccine coverage on the probabilities of developing SCT (Fig. 4B). We found that up to 50% coverage of the adult population with a vaccine with 70% effectiveness has moderate effect on risk of SCT from alpha, with 70% coverage needed to reduce the risk to a similar level to that of the ancestral strain in a naive population. For a vaccine with 90% effectiveness, 50% coverage achieves a similar risk reduction (Supplementary Fig. S7). In stark contrast to these results, in the scenarios with delta clusters, we found that 70% coverage only moderately reduces the risk of SCT relative to a naive population, and would still leave the Queensland population in a highly susceptible state with a risk of SCT of ∼ 80% for small clusters (∼ 4). For a vaccine with 90% effectiveness, 70% coverage achieves a modest additional reduction to ∼ 70% for the same cluster size (Supplementary Fig. S7). Our vaccine effectiveness values cover the range encompassing values reported in very recent studies of effectiveness of vaccines against the alpha variant [26] and delta variant [27, 28].

## 4. Discussion

We tailored an agent-based model of COVID-19 to Queensland, Australia. The calibrated model reproduces the 2020 wave of locally-acquired cases, and was used to predict epidemic scenarios in the face of more transmissible variants, in particular the alpha (B.1.1.7) and delta (B.1.617.2) variants. We found that for the same policy settings, a 70% increase in transmissibility translates to an up to 3-fold increase in the chance of developing SCT, while a 150% increase in transmissibility translates to an over 4-fold increase in the chance of developing SCT. We also found that a larger fraction of clusters of the ancestral strain die off, after crossing the SCT threshold (3-12%), compared to the fraction of clusters of alpha and delta variants that die after crossing SCT (¡1%) over the simulated period (60 days). We found that increases in SCT risk can be mitigated by a sustained increase in testing and subsequent Q&I, but the required increase in testing (10-fold) to mitigate risk of SCT far exceeds the maximum number of daily tests ever performed in Queensland between the period March 2020-August 2021(∼ 52,000 tests per day). Nevertheless, our results support a strong role for Q&I, in addition to border restrictions and hotel quarantine, in reducing the spread of COVID-19 in low community transmission environments [29]. This is further supported by the Queensland experience in which outbreaks of COVID-19 have been rare, small, and effectively contained.

The rarity of prolonged outbreaks in Queensland (before December 2021) also justifies the threshold used to quantify the probability of regimes with SCT, that could lead to larger outbreaks. In a global context where India counted over 400,000 daily diagnoses in the middle of May 2021, Argentina over 30,000, and Taiwan exceeded 700 daily diagnoses [30], our thresholds are, to say the least, conservative. However, in this Queensland-specific context, these thresholds are a lenient estimate of a policy change trigger under the assumption of a largely susceptible population, noting that appropriate thresholds may change as vaccination coverage increases. Our choice of threshold is supported by 3-day lockdowns in Queensland (January and March 2021), which were triggered by the detection of a single locally-acquired case.

Our vaccination results show that the large risk of developing SCT due to alpha and delta infections, together with the high vaccination effectiveness and coverage required to lower this risk to the levels of the ancestral strain, suggests that our current “new normal”, with approximately 50% of the eligible population in Queensland fully vaccinated as of early October 2021 [31], does not leave a substantial margin to sustain a zero community transmission regime if a moderate (let alone large) number of infections with the delta or newer highly-transmissible variant of concern leaks into the community. Our model calibration suggests Queensland had a lower transmissibility than previously found for New South Wales and Victoria [15, 16]. Several factors may contribute to this reduced transmissibility. These include: (i) fitting the model to diagnoses during a period with very limited testing; (ii) behavioural changes due to an increasing volume of news regarding outbreaks overseas; (iii) the fitting process reducing overall transmissibility to account for overall smaller spread of COVID-19 in Queensland than New South Wales and Victoria; (iv) having a lower population density and different mobility patterns around and between cities [32]; (v) a smaller manufacturing industry (17% of Australia’s output, compared to 28% and 32% for Victoria and New South Wales respectively), which has been associated with regional heterogeneity in outbreaks in Italy, potentially due to the intensity and duration of interactions between workers [33]; and (vi) a different latitude/climate [34].

We modelled Queensland without considering geographical effects explicitly, as has been done successfully to model COVID-19 in New South Wales and Victoria. This approximation may lead to underestimates in transmissibility within the denser population centres, given that Queensland’s population is geographically dispersed (just under 50% of Queensland’s population lives in Brisbane, compared to 66% of New South Wales in Sydney and 77% of Victoria in Melbourne). We also calibrated the model to locally-acquired case numbers rather than total cases, a large proportion of which were in hotel quarantine. Although we have not explicitly modelled the hotel quarantine system, our modelling of seeded community infections after without recorded cases of locally-acquired infections is designed, specifically, to simulate the consequences of leaks from hotel quarantine or cross-border importations (state or international). For our vaccination scenarios, we modelled an overall vaccine effectiveness that reduced the risk of infection, thus reducing transmission, for a range of vaccine coverage values (i.e., proportion of the eligible population that has received a full vaccine treatment), including the percentage of fully vaccinated people during the first week of October 2021 (https://covidlive.com.au/qld – accessed September 29th 2021). We used this simplified approach rather than explicitly modelling partial vaccination and the staged rollout because it enabled us to have quick overview of the different vaccination coverage requirements, depending on the dominant variant that triggered outbreaks. Future work modelling partial and full vaccination will incorporate very recent real-world evidence on how effective multi-dose vaccines are against the Delta variant, and how their effectiveness decreases over time after people have received either one or two doses [35]. This modelling work will be important to inform responses should any new variant emerge that falls outside the effective immunisation scope of currently available vaccines, and to determine when booster doses should delivered, as well as to plan the longer term strategy of re-opening international borders.

## Data Availability

We received the data for this project from Queensland Health following data custodian approval. The authors can provide details of this approach for others who want to gain access to the same or similar data.

https://github.com/brain-modelling-group/covasim-qld-model

## Author Contributions

PSL, NJS, and JAR designed research; PSL conducted simulations, calibration, analysis, figure generation and wrote the first draft of the paper; RMS, RGA, and CCK developed Covasim and provided feedback and resources to perform calibration and analysis; SBL provided data and data administration resources; PSL, NJS, JCP, and JAR acquired funding. All authors interpreted results, reviewed and edited the final version of the paper.

## Additional Information

All authors declare they have no conflict of interest.

