## Supplementary Information for "Risk of sustained SARS-CoV-2 transmission in Queensland, Australia"

### Supplementary Information:
### Risk of sustained SARS-CoV-2 transmission in Queensland, Australia

Paula Sanz-Leon*, Nathan J. Stevenson, Robyn M. Stuart, Romesh G. Abeysuriya, James C. Pang, Stephen B. Lambert, Cliff C. Kerr, James A. Roberts.

#### Appendix A. Supplementary Tables

Supplementary Table S1 summarizes the references for the most relevant categories of parameters.

| **Parameter category** | **reference/value** |
|---|---|
| Covasim version | 1.7.6 [14] |
| Age-specific susceptibility, disease progression, and mortality risks | Table 2 [16] |
| Population subsets included in each contact network | Covasim default values [14] |
| Mean number of contacts per person in settings or during activities | Table 4 [16] |
| Relative transmissibility of contact networks | as per NSW model [15] |
| Quarantine and Isolation factor | Table 7 [16] |
| Probability of successful contact tracing | Table 7 [16] |

Supplementary Table S1: Summary of references for model parameters.

| **Layer** | Value used in this work | |
|---|---|---|
| Home | 1 | 1 |
| School | 2 | 0.95 |
| Work | 2 | 0.8 |
| Community | 14 | 0.05 |
| Places of worship | 5 | 0.5 |
| Professional sports | 3 | 0.8 |
| Community Sports | 3 | 0.5 |
| Entertainment | 7 | 0.1 |
| Cafe and Restaurants | 7 | 0.7 |
| Pub and Bars | 7 | 0.5 |
| Public Transport | 14 | 0.5 |
| Public Parks | 21 | 0 |
| Large Events | 21 | 0.05 |
| Social | 21 | 0.9 |

Supplementary Table S2: Average number of days that takes to trace a contact of a confirmed positive case. The second column shows the probabilities of successfully tracing a contact in each layer.

#### Appendix B. Supplementary Figures

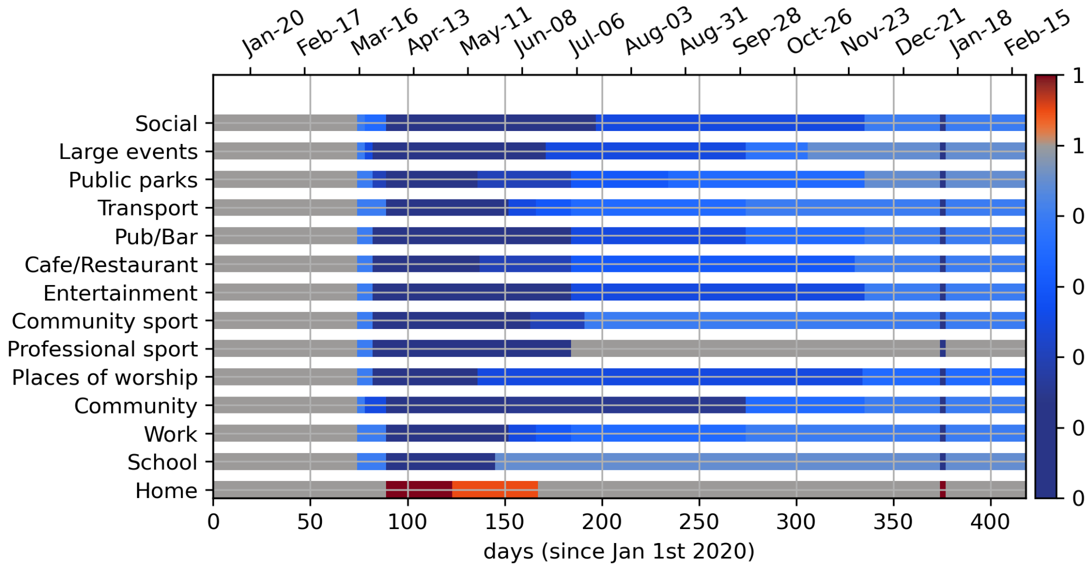


Supplementary Figure S1: Policy-based government interventions are incorporated in the model as changes in the transmissibility of each layer. In each horizontal bar, grey portions indicate the reference level transmissibility (equal to 1). Blue portions indicate the current policies have the effect of reducing the layer's transmissibility. The darker the shade of blue, the lower the effective transmissibility in that layer. Red portions indicate that the transmissibility in that layer (here, only the home layer) is larger than baseline levels because people spend more time or are in close proximity with contacts within the layer. These changes in transmissibility were approximated by looking at the changes of policies described in multiple sources [19–21].

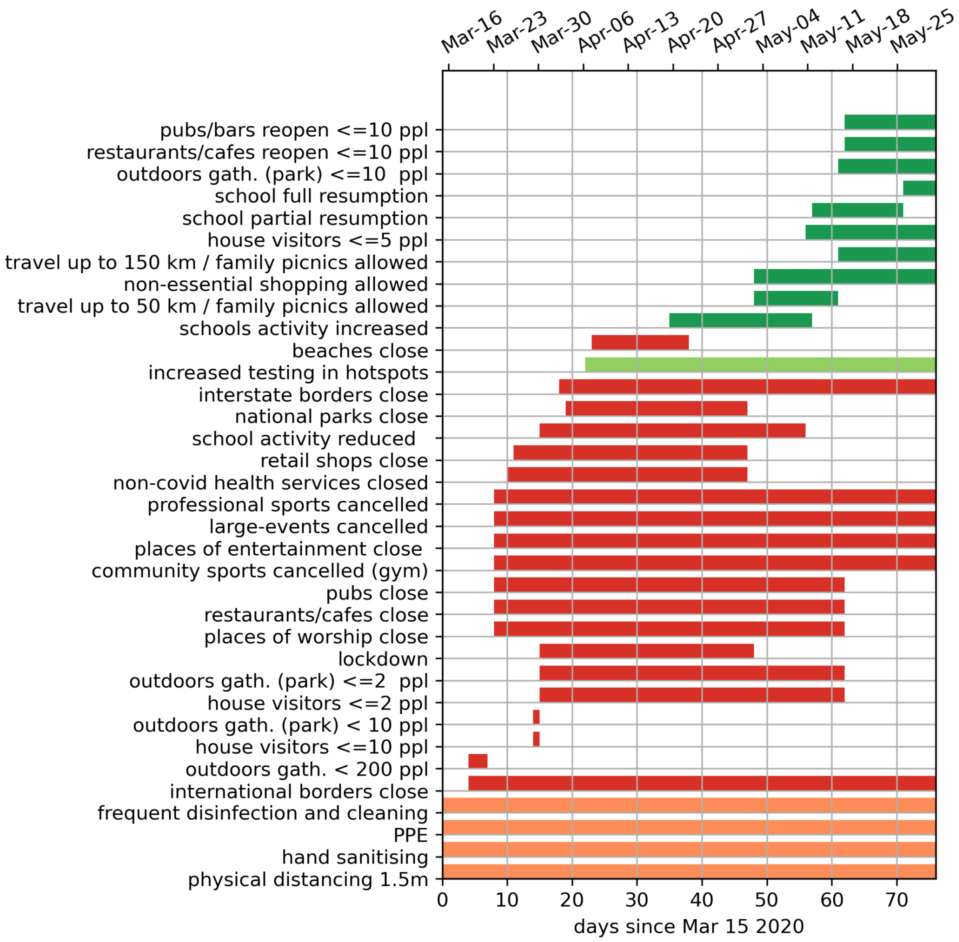


Supplementary Figure S2: Policies incorporated in the model. The colours denote four policy categories: semi-permanent public health recommendations (orange); restriction policies that reduce the spread by restricting activities of the general population (red); policies that seek to counteract the spread, such as increased testing or checking people are respecting home-quarantine (light green); and relaxation polices (green).

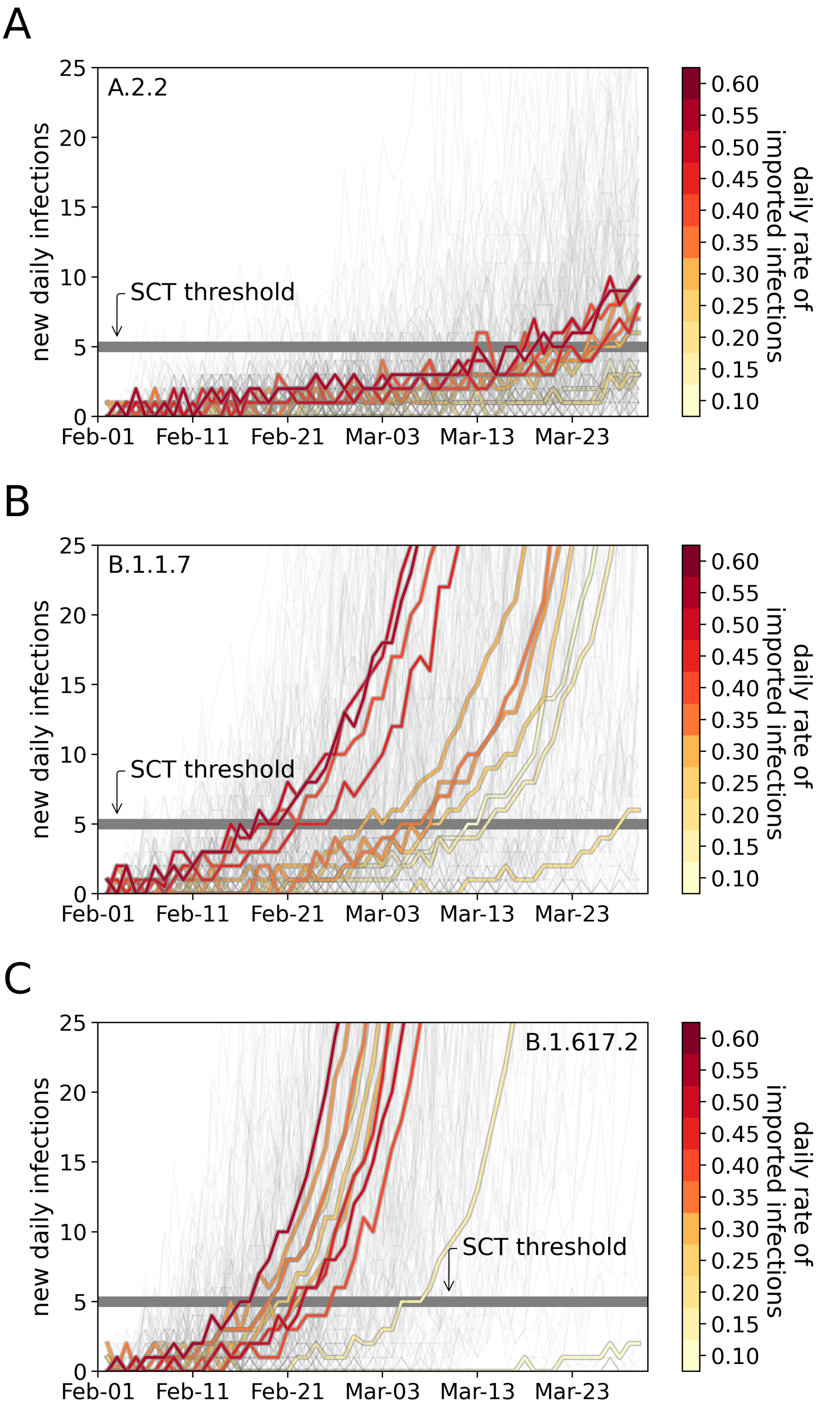


Supplementary Figure S3: Projected trajectories of new diagnoses for Poisson-seeded infections. **A**: ancestral variant. **B:** alpha variant. **C:** delta variant. Solid thick lines represent the median trajectory of new cases over 1000 runs. Light coloured lines are single model runs. The colour of each line denotes the daily rate of imported infections.

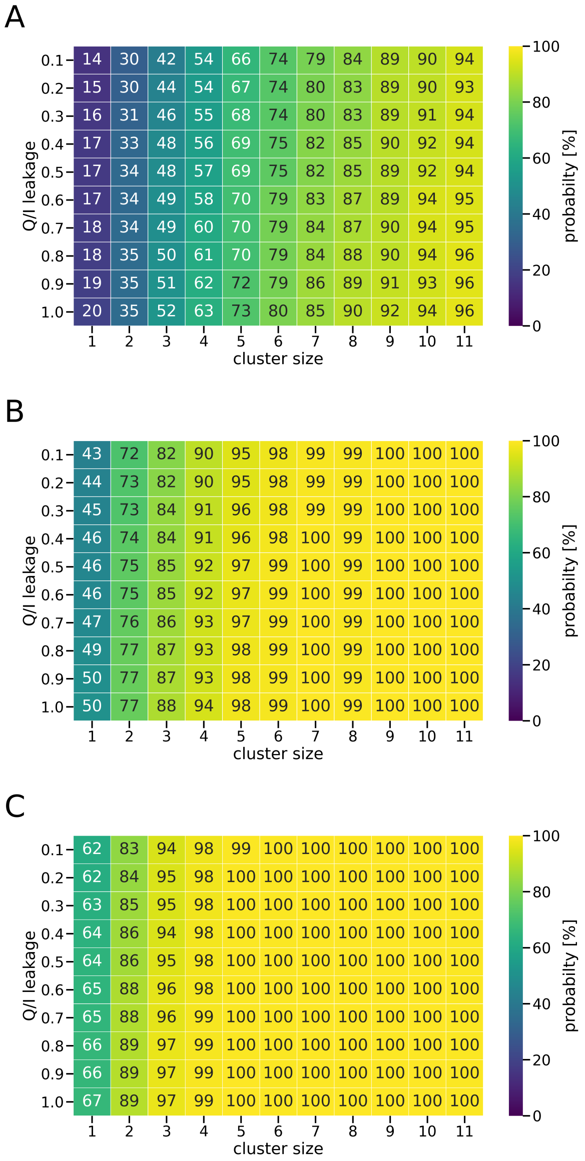


Supplementary Figure S4: Probability of regimes with sustained community transmission (SCT) for a range of Q&I factors and initial cluster sizes. **A:** ancestral variant. **B:** alpha variant. **C:** delta. Numbers in each cell and colours represent percentage (%) probabilities calculated from an ensemble of 1000 runs for each parameter set. This figure highlights the stark contrast between the risk of sustained community transmission posed by the alpha and delta variants versus the ancestral ancestral variant. For very small cluster sizes (<5), the alpha variant poses a risk of 40 percentage points more than ancestral, and the delta variant almost 50 percentage points more. In scenarios with the ancestral variant, for a cluster size of 3, Q&I leakage has a moderate effect on increasing probability of SCT from 42% (low leakage) to 52% (high leakage). However, this effect is masked in the case of the highly transmissible variants. That is, for moderate to large cluster sizes (> 7 seeded infections), community transmission is ongoing by the time the infected agents are detected and isolated, and their contacts traced and quarantined. This does not mean that Q&I interventions are not necessary but, put simply, if testing is limited, they may not be sufficient to stop SCT triggered by a small cluster of agents infected with a high-transmission variant. However, low Q&I leakage will have a crucial effect on slowing down the spread that could lead to an outbreak.

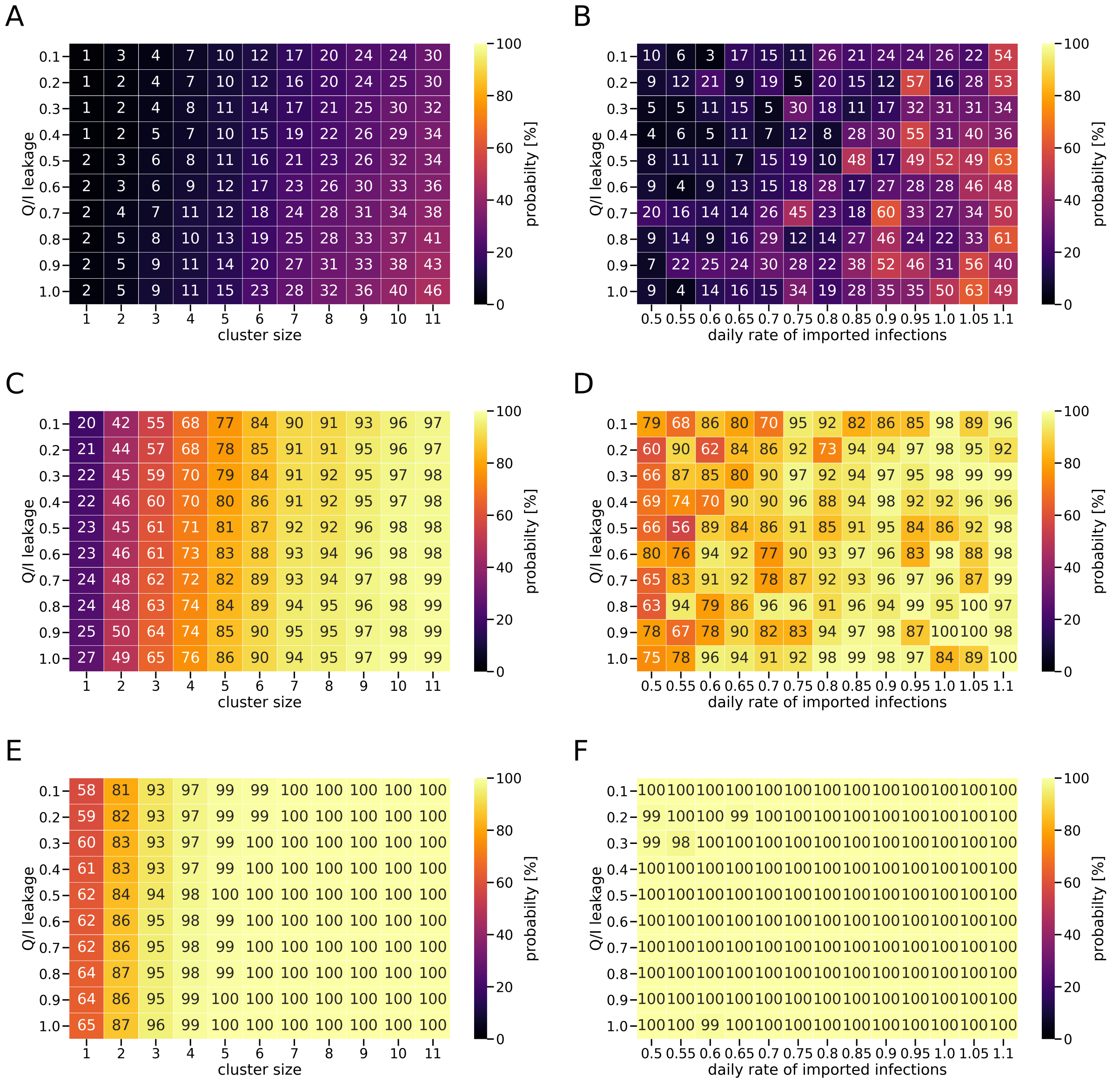


Supplementary Figure S5: Probability of detecting regimes with sustained community transmission (DSCT). **A, C and E**: Scenarios with cluster-seeded infections and ancestral, alpha and delta variants, respectively. **B, D, F**: Scenarios with Poisson-seeded infections and ancestral, alpha and delta variants, respectively. Colours and numbers represent percentage (%) probabilities calculated with respect to an ensemble of 1000 runs for each parameter set.

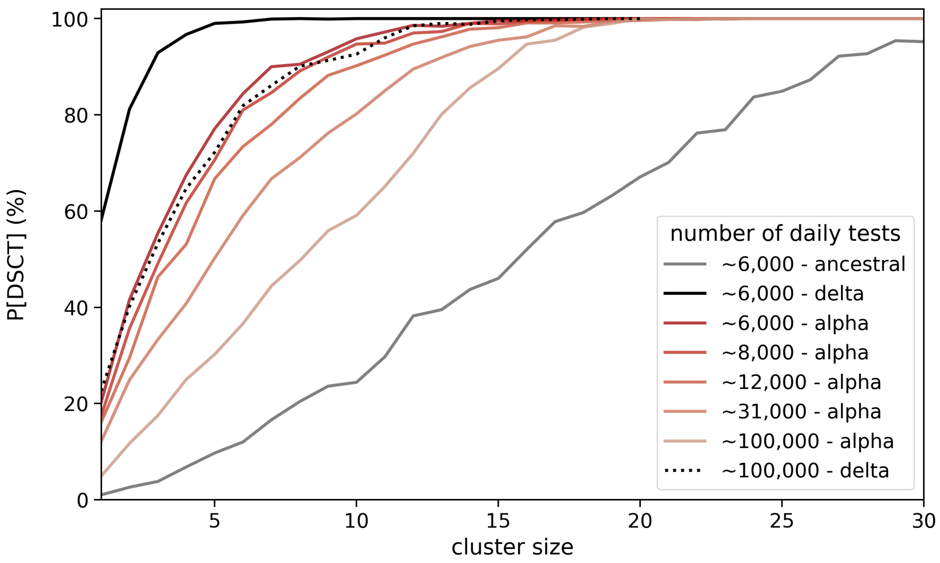


Supplementary Figure S6: Probability of detecting sustained community transmission (DSCT) for multiple cluster sizes of the highly transmissible variants. The dark grey line represents DSCT probabilities for ancestral and $\sim$ 6000 daily tests. The black line represents DSCT probabilities for B.1.167.2 and $\sim$ 6000 daily tests. The dotted black line represents DSCT probabilities for B.1.167.2 and $\sim$ 100,000 daily tests. Red-shaded lines are DSCT probabilities for the alpha variant and multiple levels of testing. All cases have a low Q&I leakage (<10%).

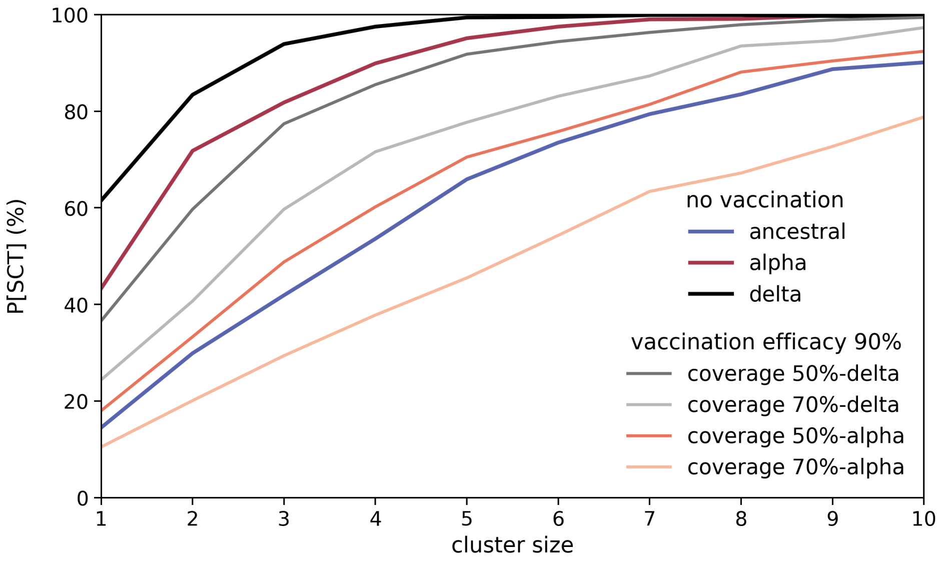


Supplementary Figure S7: Probabilities of SCT for multiple combinations of variant and vaccine coverage, for a vaccine with 90% effectiveness against transmission. The black, dark red and dark blue lines represent SCT probabilities for delta, alpha, and ancestral respectively, assuming $\sim$ 6000 daily tests, and no vaccination. Solid lines are SCT probabilities as a function of vaccination coverage, that is the percentage of the adult population that has been fully vaccinated (also known as vaccine coverage). Red line: 50% vaccinated and delta; light red line 70% vaccinated and delta; light blue line 50% vaccinated and alpha; and, blue line 70% vaccinated and alpha. In the scenarios with vaccination, the cluster size represents the number of infections seeded on the first day of the simulated period. All cases have a low Q&I leakage (<10%).
